# *CALM1, CALM2,* and *CALM3* expression and translation efficiency provide insight into the severity of calmodulinopathy

**DOI:** 10.1101/2025.05.15.25327594

**Authors:** Steffan Noe Niikanoff Christiansen, Stine Bøttcher Jacobsen, Jeppe Dyrberg Andersen, Ya Cui, Wei Li, Lia Crotti, Carla Spazzolini, Peter J. Schwartz, Mette Nyegaard, Michael Toft Overgaard

## Abstract

**Background:** Missense variants in the *CALM1, CALM2,* and *CALM3* genes cause calmodulinopathy, an ultra-rare spectrum of clinical manifestations, including ventricular arrhythmias and sudden cardiac death. The three genes encode an identical calmodulin protein. However, little is known about the potential different clinical consequences of protein-altering variants in each of the three genes.

**Methods:** We compared variants from the International Calmodulinopathy Registry with population data from the Genome Aggregation Database (gnomAD). RNA sequencing data from the Genotype-Tissue Expression (GTEx) project was used to investigate the relative expression of *CALM1, CALM2,* and *CALM3* among 49 different human tissues. We used ribosome profiling data of left ventricle tissue to estimate the percentage of calmodulin that could be attributed to each of *CALM1*, *CALM2*, and *CALM3*.

**Results:** The percentage of carriers with cardiac events was different among carriers with missense variants in *CALM1, CALM2,* and *CALM3* (*p* = 0.004). Observed-to-expected ratios in population data were higher in *CALM3* (0.29) in comparison to *CALM2* (0.20) and especially, *CALM1* (0.11) suggesting differential selection against missense variants in the three genes. RNA sequencing data revealed that *CALM3* was less expressed than *CALM1* and *CALM2* in 39 of the 49 investigated tissues. Finally, the contribution to the total calmodulin amount in the left ventricle differed significantly among *CALM1*, *CALM2*, and *CALM3* (*p* < 2×10^-16^), with estimated contributions of 45%, 44%, and 11%, respectively.

**Conclusions:** Our findings reveal a four-fold greater contribution of *CALM1* and *CALM2* to the total calmodulin levels in the human heart compared to *CALM3*, suggesting why missense variants in *CALM3* are associated with a less severe cardiac phenotype than those in *CALM1* and *CALM2*.

## Background

DNA variants that change the protein sequence of any of the calmodulin-encoding genes (CALM1, CALM2, and CALM3) cause calmodulinopathy, an ultra-rare disorder associated with a broad spectrum of clinical manifestations, including neurodevelopmental features, ventricular arrhythmias, and sudden cardiac death in childhood and adolescence (Crotti et al. 2019, 2023). While some missense variants are observed to follow a dominant inheritance pattern characterized by variable expressivity of symptoms among the carriers across multiple generations (Nyegaard et al. 2012; Kato et al. 2022), approximately 80% of the index cases are assumed to be de novo (Crotti et al. 2023).

The *CALM1*, *CALM2*, and *CALM3* genes are highly conserved, with the calmodulin amino acid sequence being identical and evolutionarily conserved across all vertebrates (Friedberg and Rhoads 2001). Due to the genetic redundancy of *CALM1*, *CALM2*, and *CALM3,* it has been shown that reduced gene expression by CRISPR interference was correcting the pathogenic phenotype in induced pluripotent stem cell-derived cardiomyocytes (Limpitikul et al. 2017). Similarly, Bortolin et al. demonstrated that arrhythmic characteristics caused by *CALM1* missense variants could be alleviated by *CALM1* depletion through specific antisense oligonucleotides (Bortolin et al. 2024). Furthermore, a combined suppression and replacement therapy can be used to deplete the expression of all three calmodulin coding genes and replace it with a fully functional *CALM1* copy to shorten pathologically prolonged action potential durations in cardiomyocyte models (Hamrick et al. 2024).

Given that arrhythmias can be alleviated by depletion in model systems, one may consider the evolutionary cause of the genetic redundancy of *CALM1*, *CALM2*, and *CALM3*. Three genes may triple the risk of *de novo* mutations that causes lethal arrhythmias. If *CALM1*, *CALM2*, and *CALM3* were simply executing the same roles, it would be expected that risk profiles of calmodulinopathy patients would be identical irrespective of the gene affected. Similarly, the same negative selection would be expected among variant carriers.

To improve the risk stratification of calmodulinopathy, we examined the genetic variation, gene expression, and translational efficiency of *CALM1*, *CALM2*, and *CALM3*. First, we assessed the pathogenicity of the missense variants by comparing data from calmodulinopathy patients in the International Calmodulinopathy Registry (Crotti et al. 2023) with population data from the Genome Aggregation Database (gnomAD) (Karczewski et al. 2020). Next, we compared gene expression among *CALM1*, *CALM2*, and *CALM3* in 49 tissues using data from the Genotype-Tissue Expression (GTEx) project (Lonsdale et al. 2013). Last, we examined the translational efficiency of *CALM1*, *CALM2*, and *CALM3* in left ventricular tissue and integrated these findings with the GTEx data to evaluate the relative contribution of each gene to the shared pool of calmodulin protein.

## Materials and Methods

### *CALM1*, *CALM2*, and *CALM3* missense variants in the International Calmodulinopathy Registry and the Genome Aggregation Database

We used data from two cohorts to investigate the potential selection pressure on variants in *CALM1*, *CALM2*, or *CALM3*. The International Calmodulinopathy Registry is a multicentre clinical observational registry that was established in 2015 to recruit patients carrying a pathogenic variant in *CALM1*, *CALM2*, or *CALM3* independent of their phenotype (Crotti et al. 2019). Clinical characteristics of 140 subjects with missense variants in *CALM1*, *CALM2*, or *CALM3* were obtained from the most recent version of the International Calmodulinopathy Registry (Crotti et al. 2023).

The Genome Aggregation Database (gnomAD) is a collection of data from large-scale sequencing projects. The most recent release (v4) includes data from 730,947 exomes and 76,215 genomes. We used the constraint metrics file (gnomad.v4.1.constraint_metrics.tsv) to get the observed-to-expected ratios of missense variants and predicted loss-of-function variants. The expected number of mutations per gene was estimated based on a mutational model as previously described (Karczewski et al. 2020). The data was filtered based on the “mane_select” variable to obtain a single representative transcript for each gene as previously described (Morales et al. 2022).

### RNA-sequencing data from the Genotype-Tissue Expression project

RNA sequencing data was obtained from the GTEx project, which is a database and public resource that has been established with the primary goal of studying gene expression among human tissues (Lonsdale et al. 2013). The samples were collected from non-diseased tissues of donors of ≥ 21 and ≤ 70 years of age (Carithers et al. 2015).

We used long-read RNA sequencing data obtained from GTEx v9 (Glinos et al. 2022) to determine which *CALM1*, *CALM2*, and *CALM3* transcripts that were expressed in human tissues. The short-read RNA sequencing data was obtained from GTEx v8 (Consortium 2020) and used to determine the relative gene expression among *CALM1*, *CALM2*, and *CALM3* in multiple tissues. The mapped read counts were generated according to the procedures described in the GTEx portal (https://gtexportal.org/home/methods). The transcript annotations were performed using the GENCODE (v26) annotation file.

### RNA-sequencing and ribosome profiling data from left ventricle tissue

RNA sequencing and ribosomal profiling data was obtained from a previous investigation of left ventricle tissue (Heesch et al. 2019). In brief, the study included data from 22 female and 58 male subjects with an average age of 43.6 years ranging from 1 to 77 years. The majority of the samples were obtained during surgical procedures of individuals affected by dilated cardiomyopathy (n=65) while the remaining samples were obtained from unused donor hearts, autopsy after trauma or from valve sparing aortic root replacement.

Ribosomal profiling enables investigation of the translating ribosomes by sequencing of the short mRNA fragments that are protected from nuclease degradation by the ribosomes (Ingolia et al. 2009). In the previous processing of the data, the reads from RNA sequencing and ribosomal profiling were trimmed to the same length and processed with the exact same settings for the analyses of the two types of data (Heesch et al. 2019). The coding sequence (CDS) mapped reads were downloaded from https://shiny.mdc-berlin.de/cardiac-translatome/.

### Data analysis

The data analysis was performed in the R statistical environment (version 4.4.1) using the tidyverse (version 2.0.0) (Wickham et al. 2019) and patchwork (version 1.3.0) (Pedersen 2024) packages. The chi-squared tests were conducted with the *chisq.test* function. The Wilcoxon signed rank test was conducted with the *wilcox.test(paired = TRUE, alternative = “two.sided”)* function. Spearman’s correlation coefficients, ρ, were calculated with the *cor.test(method = “spearman”)* function. Kruskal-Wallis rank sum test was calculated with the kruskal.test() function. The test results were only considered significant if the *p*-values were lower than the significance threshold, 0.05, divided by the number of tests.

### Estimation of 3’ untranslated region usage

The percentage of distal poly(A) site usage index (PDUI) values were calculated for the GTEx short-read sequencing data as previously described (Cui et al. 2021). The approach was based on the DaPars v.2 framework (Feng et al. 2018; Li et al. 2021). In brief, based on multiple RNA sequencing samples, the location of proximal alternative polyadenylation sites is estimated together with expression levels of the short and long 3’-untranslated regions (UTR). The output is the alternative polyadenylation usage reported per gene and sample.

#### Gene expression

The data analysis of the GTEx short-read and long-read RNA sequencing data were based on either mapped read counts or the supplied transcript per million (TPM) values. The data were filtered to only include *CALM1*, *CALM2*, and *CALM3*. We used the three representative MANE transcripts, ENST00000356978.8 (*CALM1*), ENST00000272298.11 (*CALM2*), and ENST00000291295.13 (*CALM3*) to calculate the fraction of MANE transcripts expressed compared to the sum of TPM values among all transcripts per gene.

We did not use any conventional methods for library size corrections in the analysis of the short-read sequencing data from GTEx since our main interest was to determine the percentage expressed for each of the calmodulin coding genes rather than an absolute value of expression. The percentage of counts per calmodulin coding gene per sample was calculated as shown below:

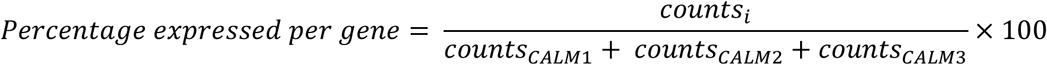

where *counts_i_*, corresponds to the counts of either *CALM1*, *CALM2*, or *CALM3*.

To account for the different gene lengths of *CALM1*, *CALM2*, and *CALM3*, within-sample comparisons among the three genes was performed with read counts adjusted for gene lengths as shown below:

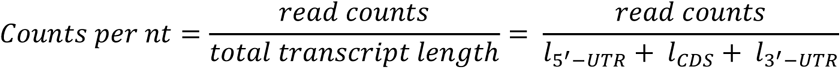

where *l_5’-UTR_*, *l_-CDS_*, and *l_3’-UTR_* correspond to the length (nt) of the 5’-UTR, CDS, and the 3’-UTR, respectively. To account for the potential underestimation of gene expression for samples with little or no 3’-UTR usage, we included the sample-specific PDUI values in the denominator as shown in the formula below:

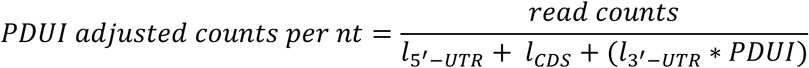

Notice that if the entire 3’-UTR is fully used, (PDUI value equal to 1), the “PDUI adjusted counts per nt” is identical to the “counts per nt” measure. For each sample, we calculated the percentage expressed from *CALM1*, *CALM2*, and *CALM3* (with and without PDUI adjustment).

#### Translational efficiency

The RNA-sequencing and ribosome profiling data was normalised simultaneously to enable integration of the two types of data for estimation of translational efficiency. The size factor was estimated to account for sequencing depths using the *estimateSizeFactorsForMatrix()* function from the DESeq2 R package (v.1.44.0) (Love et al. 2014). Since the CDS lengths of *CALM1*, *CALM2*, and *CALM3* are identical and the reads were only mapped to the CDS, we did not adjust the RNA-sequencing and ribosome profiling data for gene length. The translational efficiency was estimated for each gene per sample by taking the ratio of ribosome profiling counts over RNA-sequencing counts as previously described (Chothani et al. 2019; Heesch et al. 2019).

To obtain an estimate of the percentage of calmodulin that is translated from each of *CALM1*, *CALM2*, and *CALM3* in left ventricle tissue, we multiplied the “PDUI adjusted counts per nt” obtained from GTEx v8 with the median translational efficiency for each of the genes.

## Results

### Different severity of DNA variants in *CALM1*, *CALM2*, and *CALM3*

The calmodulin protein is encoded by three different genes, *CALM1*, *CALM2*, and *CALM3* (Fig. 1A). To investigate the potentially different clinical characteristics of DNA variants in *CALM1*, *CALM2*, and *CALM3*, we used the data from 140 individuals with a missense variant in the International Calmodulinopathy Registry (Crotti et al. 2023). There was a significant association between the calmodulin encoding genes and the fraction of individuals with any cardiac event (Chi-squared test, *p* = 0.004). As shown in Fig 1B, the percentage of carriers with a cardiac event was highest for *CALM1* (46/52, 89%) followed by *CALM2* (37/53, 70%) and *CALM3* (20/35, 57%). Predicted loss-of-function variants (incl. stop, frameshift or splice site variants) were not reported in any of *CALM1*, *CALM2*, and *CALM3* in the International Calmodulinopathy Registry.

**Figure 1.**
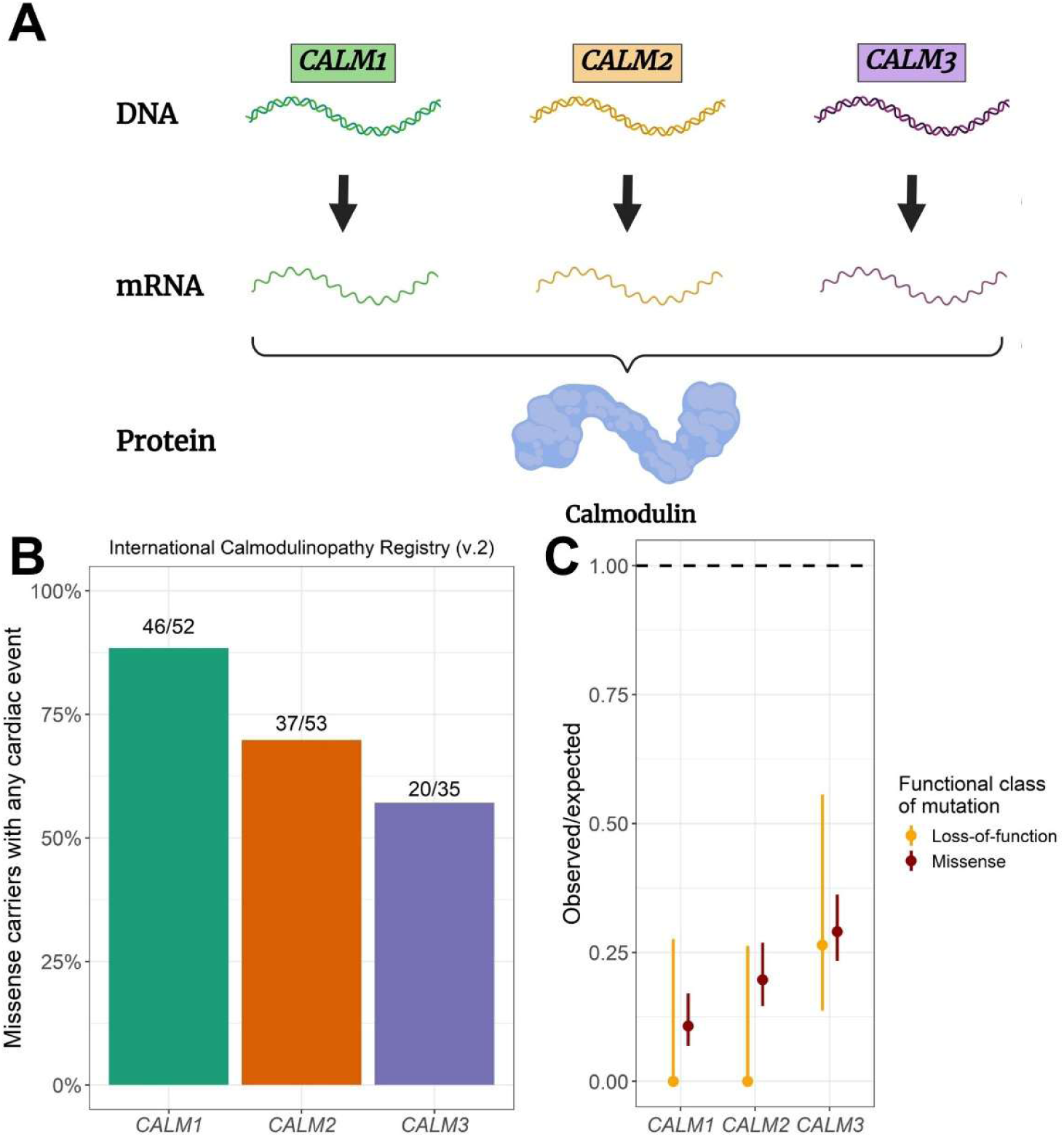
| Different clinical severity of mutations in *CALM1*, *CALM2*, and *CALM3.* A) Illustration of the unique property that the three calmodulin encoding genes contribute to the total pool of an identical calmodulin protein. B) Fraction of missense variant carriers in *CALM1*, *CALM2*, and *CALM3* who exhibit symptoms, based on data from the International Calmodulinopathy Registry. Any cardiac event corresponds to any of arrhythmic syncope, aborted cardiac arrest, sudden cardiac death or appropriate implantable cardioverter–defibrillator shocks. C) Observed-to-expected ratios of *CALM1*, *CALM2*, and *CALM3* variants in gnomAD together with 90% confidence intervals and categorized by functional class. The horizontal dashed line at observed-to-expected = 1 represents the expected number of variants under neutral selection.

To further assess the selection against missense variant of the DNA variants, we compared the observed-to-expected ratios of variants in gnomAD. An observed-to-expected ratio equal to 1 indicates that there is no selection against that particular type of variant while an observed-to-expected ratio close to zero indicates strong negative selection. The observed-to-expected ratios of missense variants were 0.11, 0.20, and 0.29 for *CALM1*, *CALM2*, and *CALM3*, respectively (Fig. 1C). Furthermore, the observed-to-expected ratio of predicted loss-of-function mutations was 0.27 for *CALM3* while it was 0 for both *CALM1* and *CALM2* (Fig. 1C).

### Identification of the representative transcripts of *CALM1*, *CALM2*, and *CALM3*

To investigate the different roles of the *CALM1*, *CALM2*, and *CALM3*, we compared the genetic structure of the genes with the GENCODE v26 annotation file. According to the annotation file, there were 16, 9, and 13 different transcripts for *CALM1*, *CALM2*, and *CALM3*, respectively. To assess the relevance of the different transcripts, we explored the TPM values of the GTEx long-read RNA sequencing data (Glinos et al. 2022). The analysis was conducted on 66 samples that represented 13 different tissues and 46 donors.

For each of *CALM1*, *CALM2*, and *CALM3*, we calculated the percentage of MANE transcripts relative to all transcripts across all samples. The percentage of MANE transcripts expressed among the tissues were 99.1% for *CALM1* (range: 96.8%-100%), 99.0% for *CALM2* (range: 96.2%-100%), and 98.7% for *CALM3* (range: 96.0%-100%). Due to the relatively low expression of the remaining transcripts, we excluded these from the rest of the analyses.

### Different structure and usage of the 3’ untranslated regions among *CALM1*, *CALM2*, and *CALM3*

The total exon length of *CALM1*, *CALM2*, and *CALM3* ranged from 1,302 to 4,242 nt among the MANE transcripts (Fig. 2A). The CDS lengths were identical for all three genes, consisting of 450 nt including the start and stop codon. In contrast, the length of the 3’-UTRs was 3,544 nt for *CALM1*, 694 nt for *CALM2*, and 1,640 nt for *CALM3*.

**Figure 2.**
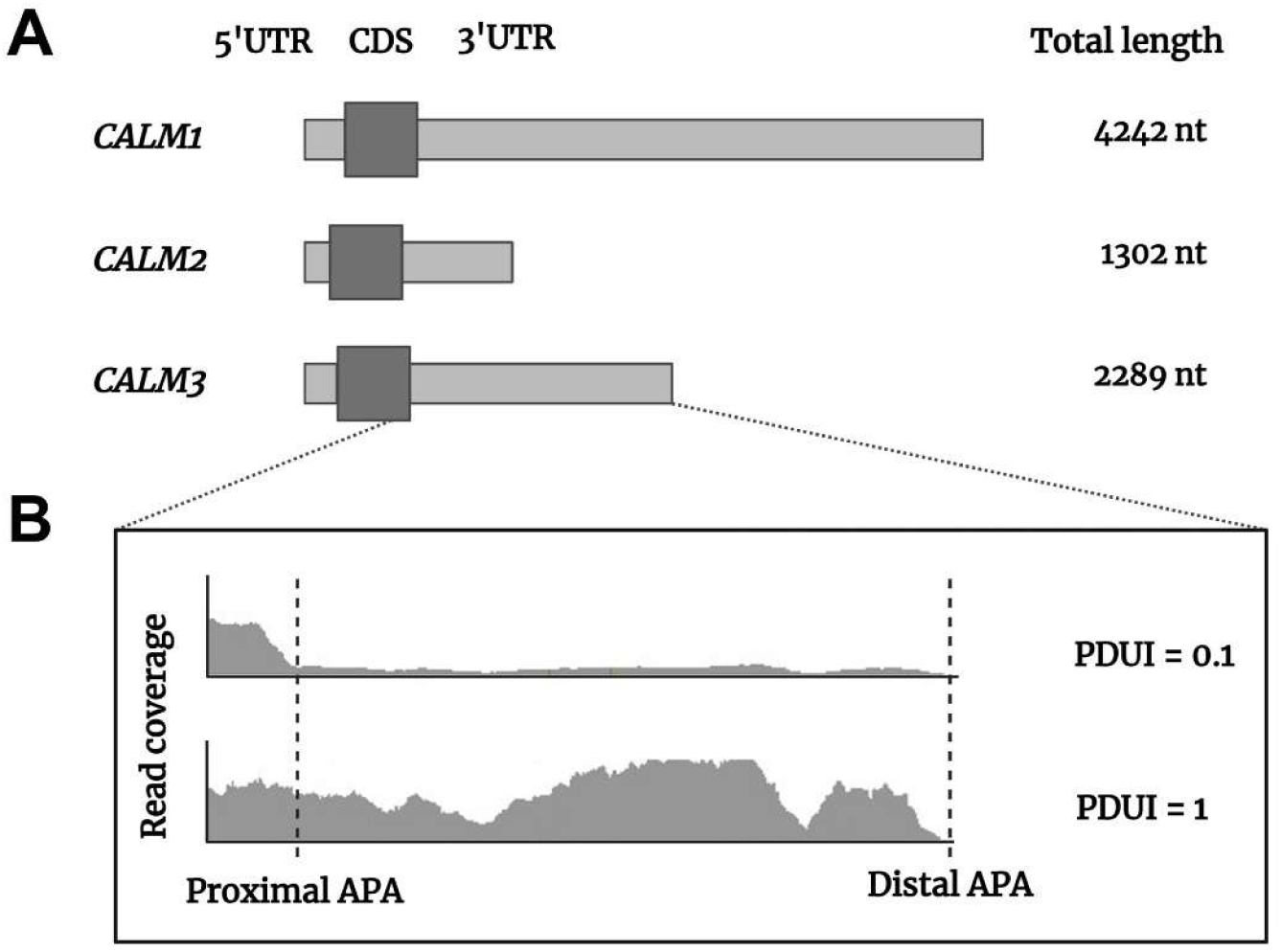
| Illustration of *CALM1*, *CALM2*, and *CALM3* transcript structures and alternative polyadenylation usage. A) The total transcript lengths (in nucleotides) are indicated to the right. The coding sequence (CDS) is shown in dark gray, while the untranslated regions (UTRs) are in light gray. B) Example of read coverage across 3’-UTR for CALM3 with proximal and distal alternative polyadenylation (APA) sites indicated by dashed lines. Percentage of distal polyA site usage index (PDUI) values represent the relative usage of distal polyadenylation sites, where higher values indicate greater distal site preference.

Commonly used measures for comparison of RNA sequencing data account for gene length as a part of the normalisation procedure (Zhao et al. 2021). However, full-length 3’-UTRs are not necessarily expressed. Furthermore, the 3’-UTR lengths differ considerably among *CALM1*, *CALM2*, and *CALM3*. For instance, the 3’-UTR length is 5.1 times longer than the CDS for *CALM1* while the 3’-UTR length is only 1.1 times longer than the CDS for *CALM2.* Therefore, the relative expression of the three genes may be inaccurately determined by conventional quantification approaches. To address this challenge, we evaluated the 3’-UTR usage by calculating the PDUI values for *CALM1*, *CALM2*, and *CALM3* (Fig. 2B). The analysis was conducted on 15,201 samples from 838 individuals for a total of 49 different tissues using the GTEx short-read RNA sequencing data as previously described (Cui et al. 2021). The PDUI values were distributed as shown in Fig. 3 with values ranging from 0.03 to 1 for *CALM1* (median PDUI: 0.32), from 0.235 to 1 for *CALM2* (median PDUI: 0.70), and from 0.12 to 1 for *CALM3* (median PDUI: 0.69).

**Figure 3.**
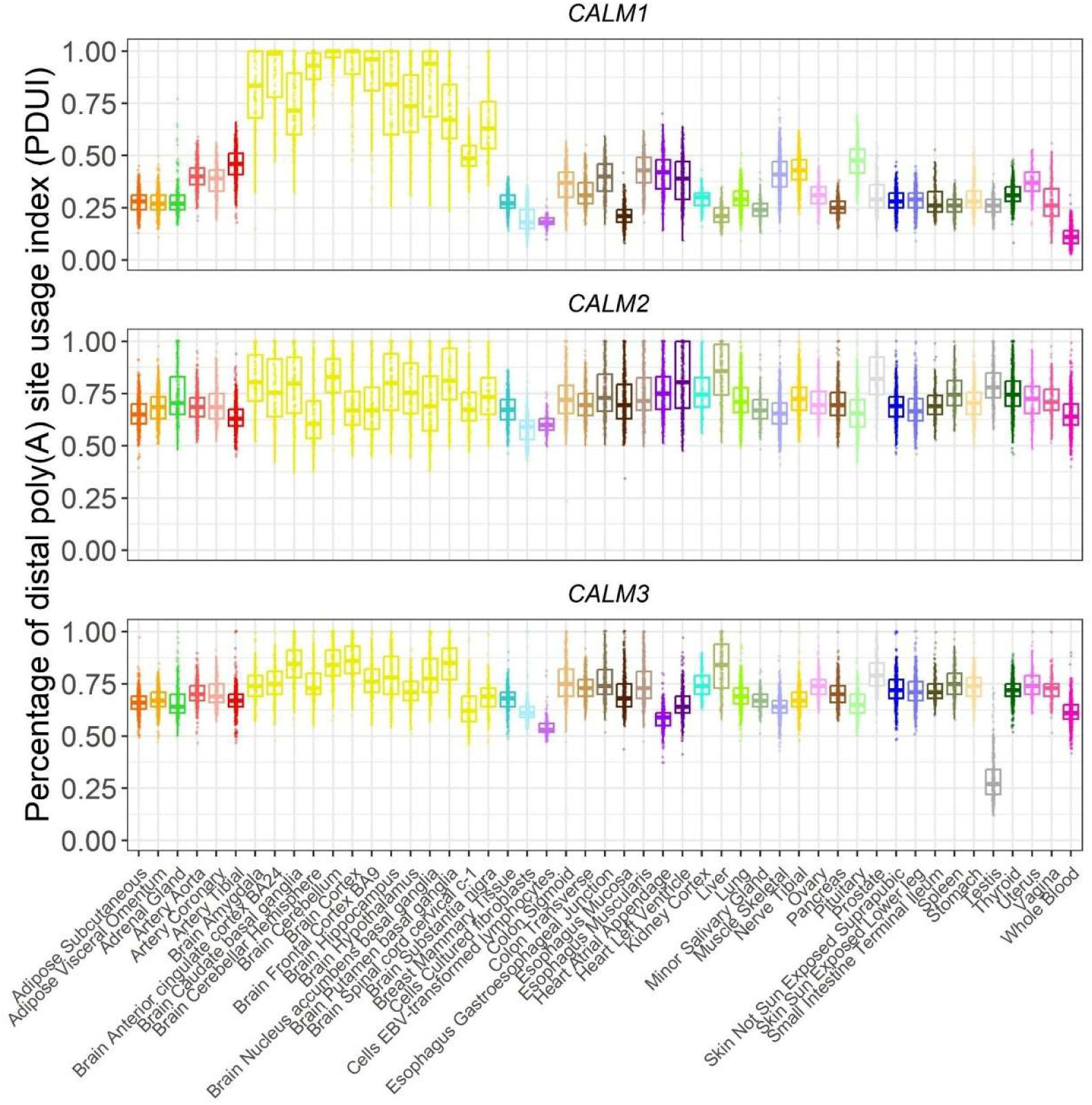
| Percentage of distal poly(A) usage index values among *CALM1, CALM2*, and *CALM3*. Box plot of percentage of distal poly(A) usage index values (PDUI) per tissue among *CALM1*, *CALM2*, and *CALM3.* Each data point corresponds to a sample-specific and gene-specific PDUI value. The color codes indicate tissue of origin and are adopted from the GTEx project.

### Adjustment of gene expression according to 3’-untranslated region usage

To illustrate the potential impact of PDUI values on gene expression quantification of the calmodulin encoding genes, we constructed an example of *CALM1* expression in two different artificial samples that differed solely in their PDUI values (Fig. 4A). In sample #1, the full-length 3’-UTR was used (PDUI = 1), resulting in no discrepancy between the values reported as “counts per nt” and “PDUI adjusted counts per nt”. In contrast, in sample #2, where the full-length 3-’UTR was not used (PDUI = 0), the “PDUI adjusted counts per nt” exhibited a 6.1-fold increase relative to the unadjusted “counts per nt.” In other words, depending on whether the PDUI value is 1 or 0, a sample with 10,000 read counts mapped to *CALM1* may represent the same number of RNA copies as another sample with only 1,639 read counts.

**Figure 4.**
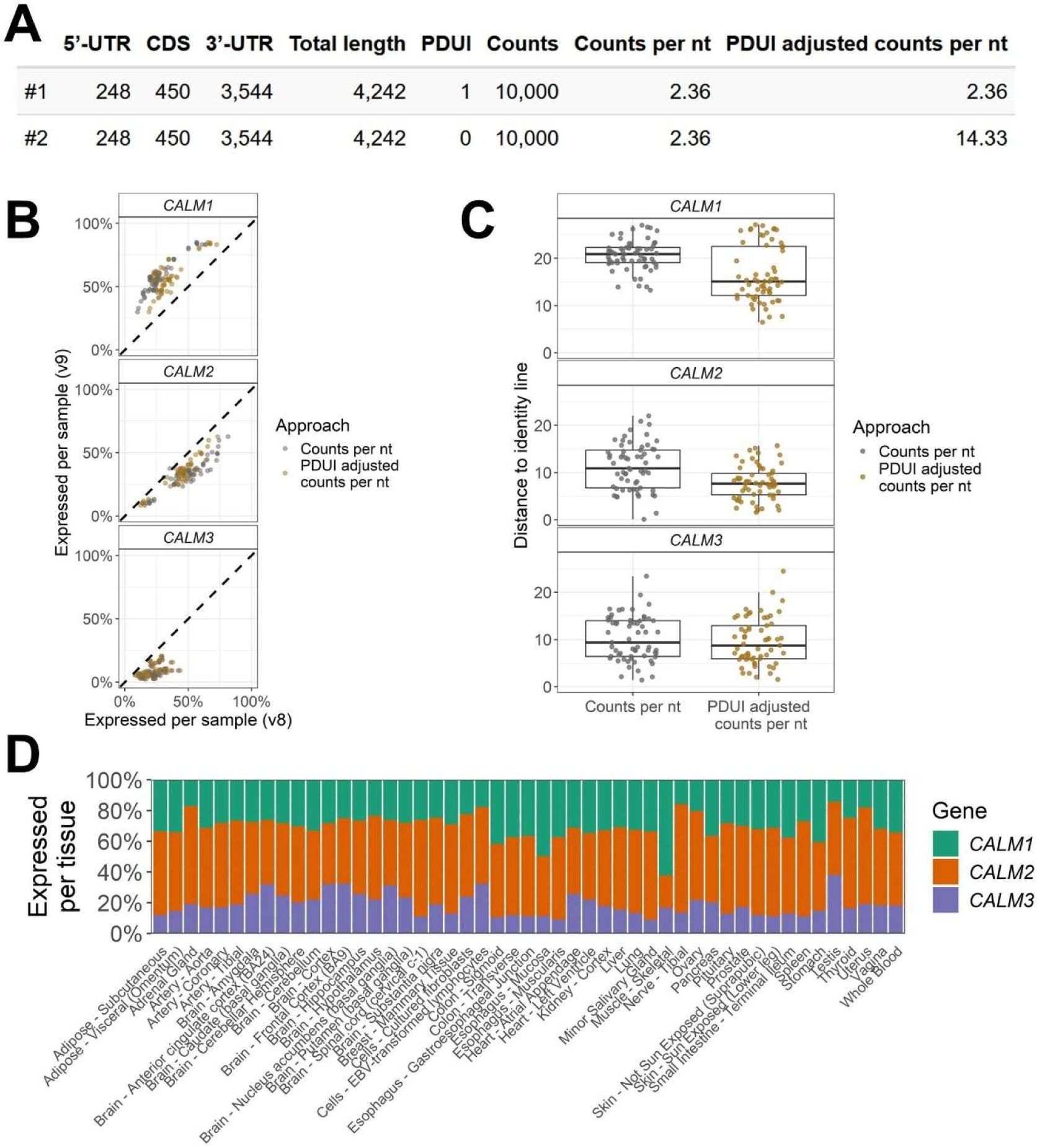
| Impact of 3’-untranslated region usage on gene expression values. A) Constructed example on the effect of 3’-untranslated region (UTR) usage estimated by percentage of distal poly(A) usage site index (PDUI) values. The lengths of the 5’-UTR, coding sequence (CDS), and 3’-UTR of *CALM1* is obtained from Gencode V26. B) Relative expression per sample for long-read sequencing data (v9) and short-read sequencing data (v8) in paired samples. Samples with identical relative expression measured with long-read and short-read sequencing data are expected to overlap the identity lines (dashed). C) Similarity measured as distance to identity line for the data shown in B). D) Percentage expressed of *CALM1*, *CALM2*, and *CALM3* per tissue among the 49 examined tissues based on PDUI adjusted counts per nt.

We tested our approach by comparing the percentage of counts that could be attributed to each of *CALM1*, *CALM2*, and *CALM3* with and without PDUI adjustment for the subset of 61 GTEx samples that were investigated with both short-read (GTEx v8) and long-read RNA sequencing (GTEx v9) (Fig. 4B). The median distances to the identity line were significantly different (Wilcoxon rank sum test) for *CALM1* (*p* = 2.81×10^-7^), *CALM2* (*p* = 4.95×10^-7^), and *CALM3* (*p* = 3.77×10^-4^). The median distance was reduced with 5.8 percentage points for *CALM1* (27.6%), 3.3 percentage points for *CALM2* (29.8%), and 0.6 percentage points for *CALM3* (6.7%) by using “PDUI adjusted counts per nt” compared to the “counts per nt” (Fig. 4C).

To demonstrate the relevance of our approach, we calculated the log_2_ transformed fold change between the percentage of counts that could be attributed to *CALM1*, *CALM2*, and *CALM3* measured as “PDUI adjusted counts per nt” and “counts per nt” for all 15,201 samples with short-read sequencing data (Fig. S1). We observed a significant difference (one-sample Wilcoxon rank sum test, *p* < 0.00034) between the two approaches for all the three genes in 46 out of 49 tissues. The median of medians for the log_2_ transformed fold change were 0.67, –0.24, and –0.12 for *CALM1*, *CALM2*, and *CALM3*, respectively (Fig. S1), which highlights that the percentage expressed is generally underestimated for *CALM1* and generally overestimated for *CALM2* and *CALM3*.

Based on the “PDUI adjusted counts per nt”*, CALM2* was the most expressed gene in 47 out of 49 tissues while *CALM1* was most expressed in the remaining two tissues, esophagus (mucosa) and skeletal muscle (Fig 4D). In contrast, *CALM1* was the least expressed gene in 10 out of 49 tissues while *CALM3* was the least expressed in the remaining 39 tissues.

### Translational efficiency of *CALM1*, *CALM2*, and *CALM3* in the left ventricle

To further examine to which extent the *CALM1*, *CALM2*, and *CALM3* genes contribute to the total calmodulin amount, we calculated the translational efficiency i.e. the number of ribosomes per transcript, based on a previous investigation using paired RNA sequencing and ribosome profiling data from left ventricle tissue (Heesch et al. 2019). The median translational efficiency varied among the three genes, corresponding to 3.01 for *CALM1*, 2.59 for *CALM2*, and 1.26 for *CALM3* (Fig. 5A).

**Figure 5.**
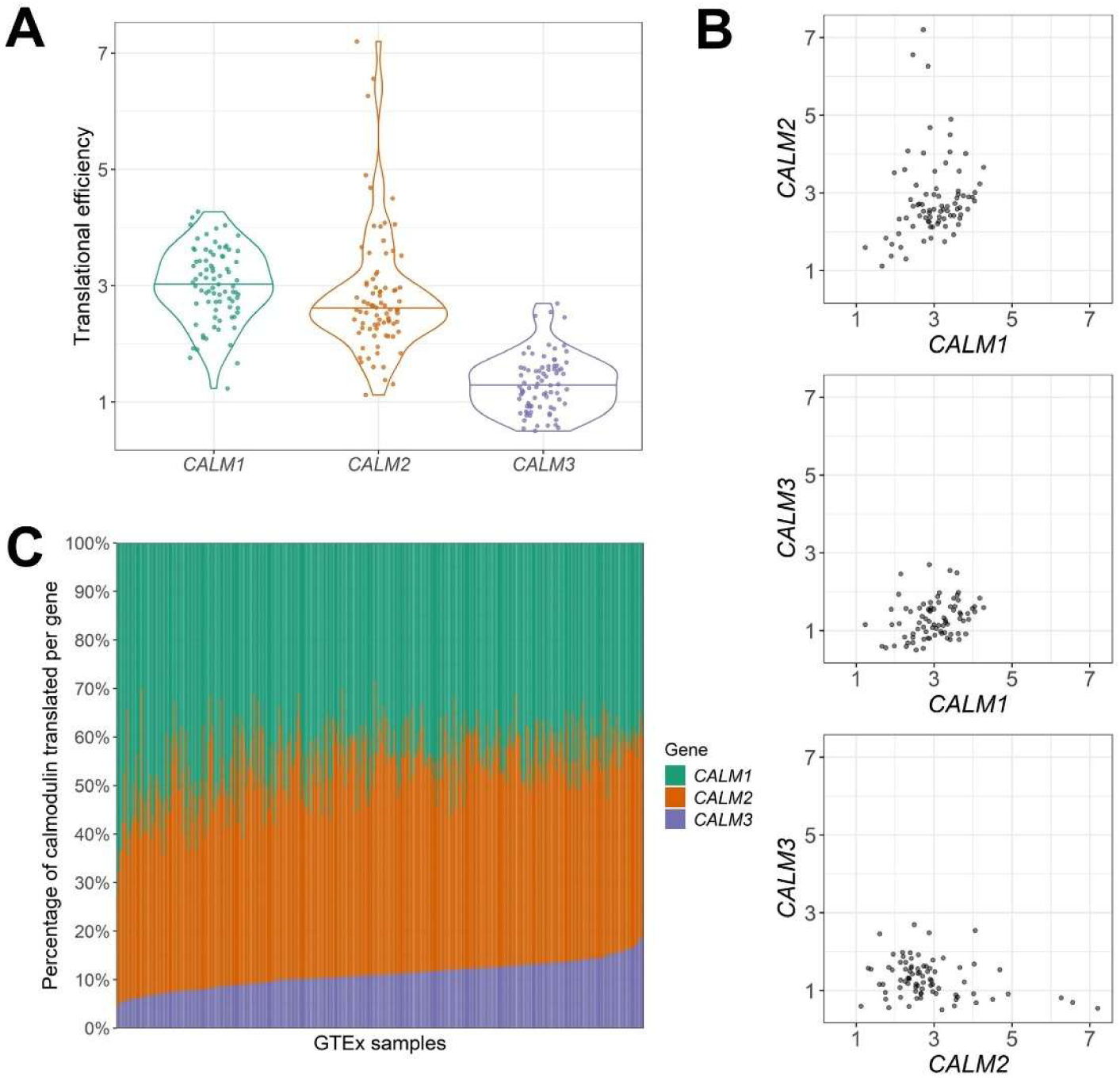
| Estimation of translation among *CALM1*, *CALM2*, and *CALM3* in left ventricle. A) Violin blots of translational efficiency per individual among *CALM1*, *CALM2*, and *CALM3* in 80 left ventricle samples. The translational efficiency was significantly different using the Wilcoxon rank sum test for *CALM1* vs *CALM2* (*p* = 2.53×10^-3^), *CALM1* vs *CALM3* (*p* = 2.79×10^-14^), and for *CALM2* vs *CALM3* (*p* = 5.51×10^-14^). B) Pairwise comparisons of translational efficiency. C) Estimated percentage of calmodulin translated per gene for left ventricle samples obtained from Genotype-Tissue Expression (GTEx) project.

Next, we conducted three pairwise comparisons of the correlations of translational efficiency among all individuals. We observed a significant positive correlation after correction for multiple testing between *CALM1* and *CALM2* (Spearman’s ρ = 0.3, *p* < 0.017), and between *CALM1* and *CALM3* (Spearman’s ρ = 0.33, *p* < 0.017) but not between *CALM2* and *CALM3* (Spearman’s ρ = –0.23, *p* = 0.037) (Fig. 5B).

Last, we integrated gene expression data from GTEx with translational efficiency values to estimate the percentage of calmodulin translated per left ventricle sample from each of *CALM1*, *CALM2*, and *CALM3* (Fig. 5C) and showed that the percentage of calmodulin translated was significantly different among *CALM1*, *CALM2*, and *CALM3* (Kruskal-Wallis rank sum test, *p* < 2×10^-16^). On average, *CALM1* accounted for 45% of the translated calmodulin (range: 28%-72%), *CALM2* accounted for 44% (range: 24%-64%), and *CALM3* accounted for 11% (range: 4%-19%). Assuming equal expression and translation of both alleles for each of the *CALM1*, *CALM2*, or *CALM3* genes, we estimated the percentage of calmodulin protein carrying a missense mutation to be, on average, 23%, 22% and 6% of the total calmodulin amount.

## Discussion

Despite encoding an identical calmodulin amino acid sequence, the negative selection and clinical characteristics vary greatly for *CALM1*, *CALM2*, and *CALM3*. In this study, we identified key differences in 3’ UTR usage, gene expression, and translational efficiency based on our multi-omics approach for addressing the genetic redundancy of *CALM1*, *CALM2*, and *CALM3*.

Risk stratification of calmodulinopathy patients often relies on the comparison of the same missense variants in one of the other two calmodulin encoding genes due to the rarity of the disease. By combining results of clinical and population investigations, we highlight that *CALM3* is generally subject to less negative selection than *CALM1* and *CALM2* (Fig. 1C) and that the cardiac events are more prevalent among *CALM2* and, especially, *CALM1* carriers (Fig. 1B).

*CALM3* was previously shown to be the most expressed calmodulin coding gene in left ventricle (Crotti et al. 2013). However, since the investigation was based on quantitative reverse transcription PCR, it is possible that different efficiencies among the primers targeted against *CALM1-3* may have caused a higher reported expression of *CALM3*. We show that *CALM3* is the least expressed calmodulin coding gene among most tissues (Fig. 4D). Furthermore, we provide the first evidence that *CALM1* and *CALM2* contribute substantially more than *CALM3* to the shared calmodulin pool in left ventricle tissue (Fig. 5C). Our coupled usage of clinical, genetic, and transcriptomic data provides a unique opportunity, even in an ultra-rare disease, to unravel the genotype-phenotype correlation to allow gene specific risk stratification which is the key to personalised clinical management.

Based on the integration of RNA sequencing and ribosome profiling data, we provide novel and unique evidence for the variable contribution from each of *CALM1*, *CALM2*, and *CALM3* to the total protein amount. For instance, the estimated *CALM3* contribution to the total calmodulin amount ranged from 4%-19% corresponding to more than a four-fold difference among the individuals. This observation is crucial considered that the clinical manifestations of calmodulinopathy may be asymptomatic or severe in the form of sudden cardiac deaths even within the same family (Kato et al. 2022). We assume that the variable dosage of mutated calmodulin compared to normal calmodulin is partly responsible for the variable spectrum of symptoms within the same families.

We comprehensively profiled the 3’-UTR usage among 49 human tissues. Consistent with previous investigations of the 3’-UTR of *CALM1* in rodents (Tushev et al. 2018; Bongmin et al. 2020), we observed increased usage of the long 3-’UTR in brain tissues compared with other tissues (Fig. 3). Our findings demonstrate the important technical implications of variable 3’-UTR usage when comparing gene expression. The quantification bias was particularly pronounced for *CALM1* due to its relatively long 3’-UTR. We anticipate that variable 3’-UTR usage may affect the reported gene expression in other studies of genes with relatively long 3’-UTRs and variable 3’-UTR usage.

Novel therapeutic approaches for calmodulinopathy rely on the depletion of the gene affected by a missense mutation (Bortolin et al. 2024; Hamrick et al. 2024). Specifically, Bortolin et al. showed that the total calmodulin levels were maintained after *CALM1* depletion due to rescue by *CALM2* and *CALM3* (Bortolin et al. 2024). Therefore, we examined the pairwise correlations of translational efficiencies among *CALM1*, *CALM2*, and *CALM3*. If *CALM1* depletion should be translationally rescued by *CALM2* or *CALM3*, a negative correlation between translational efficiencies would have been observed. However, we observed positive correlations for *CALM1* versus *CALM2* and for *CALM1* versus *CALM3* (Fig. 5B) suggesting that the genes are translationally coregulated but not compensating for each other. We identified different signatures of *CALM1*, *CALM2*, and *CALM3* in terms of 3’-UTR usage, expression levels, and translational efficiency. While depletion-based therapeutics provide a novel and promising opportunity to treat the severe symptoms of calmodulinopathy, we argue that it is important to understand the unique role of the three genes in different tissue to avoid potential adverse effects.

In this study, we focused on the cardiac features of calmodulinopathy but acknowledge that the neurodevelopmental features may also cause the observed negative selection of *CALM1*, *CALM2*, and *CALM3* mutations. Since individual-level phenotypic characteristics were unavailable from the gnomAD cohort, we were unable to investigate the fraction of carriers with asymptomatic, neurodevelopmental, or cardiac characteristics. Our study was based on bulk RNA sequencing profiles. While we gained considerable insight into the role of *CALM1*, *CALM2*, and *CALM3* using our approach, we argue that future studies should be designed to address the diversity of expression among single cells.

## Conclusion

Overall, our work suggests that *CALM1*, *CALM2*, and *CALM3* exert different roles. Although missense variants in *CALM3* are generally pathogenic, we provide the first evidence that the consequence of missense variants in *CALM3* is less severe compared to missense variants in *CALM1* and *CALM2*.

## Declarations

### Ethics approval and consent to participate

The study was approved by the Institutional Review Board of Aalborg University (AAU031-1058687). The analyses were conducted on data that were either publicly available or available upon application (dbGaP: phs000424.v8.p2). No additional data were generated for this study. The study was conducted in accordance with the Declaration of Helsinki.

### Consent for publication

Not applicable.

### Availability of data and materials

We used the constraint metrics data file (gnomad.v4.1.constraint_metrics.tsv) obtained from https://gnomad.broadinstitute.org/data. Data from the International Calmodulinopathy Registry were available from the online version of the paper published in the European Heart Journal (Crotti et al. 2023). The mapped read counts from GTEx were downloaded from the GTEx Portal. Counts from the short-read sequencing data (v8 release) was downloaded from: https://gtexportal.org/home/downloads/adult-gtex/bulk_tissue_expression. The long-read RNA sequencing data (v9 release) was downloaded from https://gtexportal.org/home/downloads/adult-gtex/long_read_data. The raw reads for the calculation of the PDUI values were obtained from dbGaP (accession number phs000424.v8.p2). The GENCODE (v26) annotation file was downloaded from: https://ftp.ebi.ac.uk/pub/databases/gencode/Gencode_human/release_26/. The RNA-sequencing and ribosome profiling data from the 80 left ventricle samples were downloaded from a web application (https://shiny.mdc-berlin.de/cardiac-translatome/) (accessed 2022.11.29).

## Funding

SNNC was supported by a research grant from the Danish Cardiovascular Academy, which is funded by the Novo Nordisk Foundation, grant number NNF20SA0067242 and the Danish Heart Foundation. MTO and PJS were supported by a Novo Nordisk Foundation Tandem Programme grant (NNF23OC0081817, The Calmodulinopathy Research Program: Unraveling the Mechanistic Link between Calmodulin Mutations and Clinical Phenotypes).

### Authors’ contributions

Conceptualisation: SNNC, MN, and MTO. Data analysis: SNNC, SBJ, YC, WL, CS, and LC. Interpretation of the results: SNNC, SBJ, JDA, CS, LC, PJS, MN, and MTO. Visualisation: SNNC. Writing and revising the manuscript: SNNC, SBJ, JDA, CS, LC, PJS, MN, and MTO. All authors read and approved the final version of the manuscript.

### Software and code

The code for the analyses will be available from GitHub (https://github.com/SteffanChristiansen).

## Data Availability

All data are available publicly available or available upon application (see section "Availability of data and materials").

## Acknowledgements

LC and PJS are proud members of the European Reference Network for Rare and Low Prevalence Complex Diseases of the Heart (ERN GUARD-Heart). The GTEx Project was supported by the Common Fund of the Office of the Director of the National Institutes of Health, and by NCI, NHGRI, NHLBI, NIDA, NIMH, and NINDS. The data for the analysis and availability is described in the “Availability of data and materials” section. Figs. 1A and 2A-B were created with BioRender.com.

## Figures

**Supplementary figure 1.**
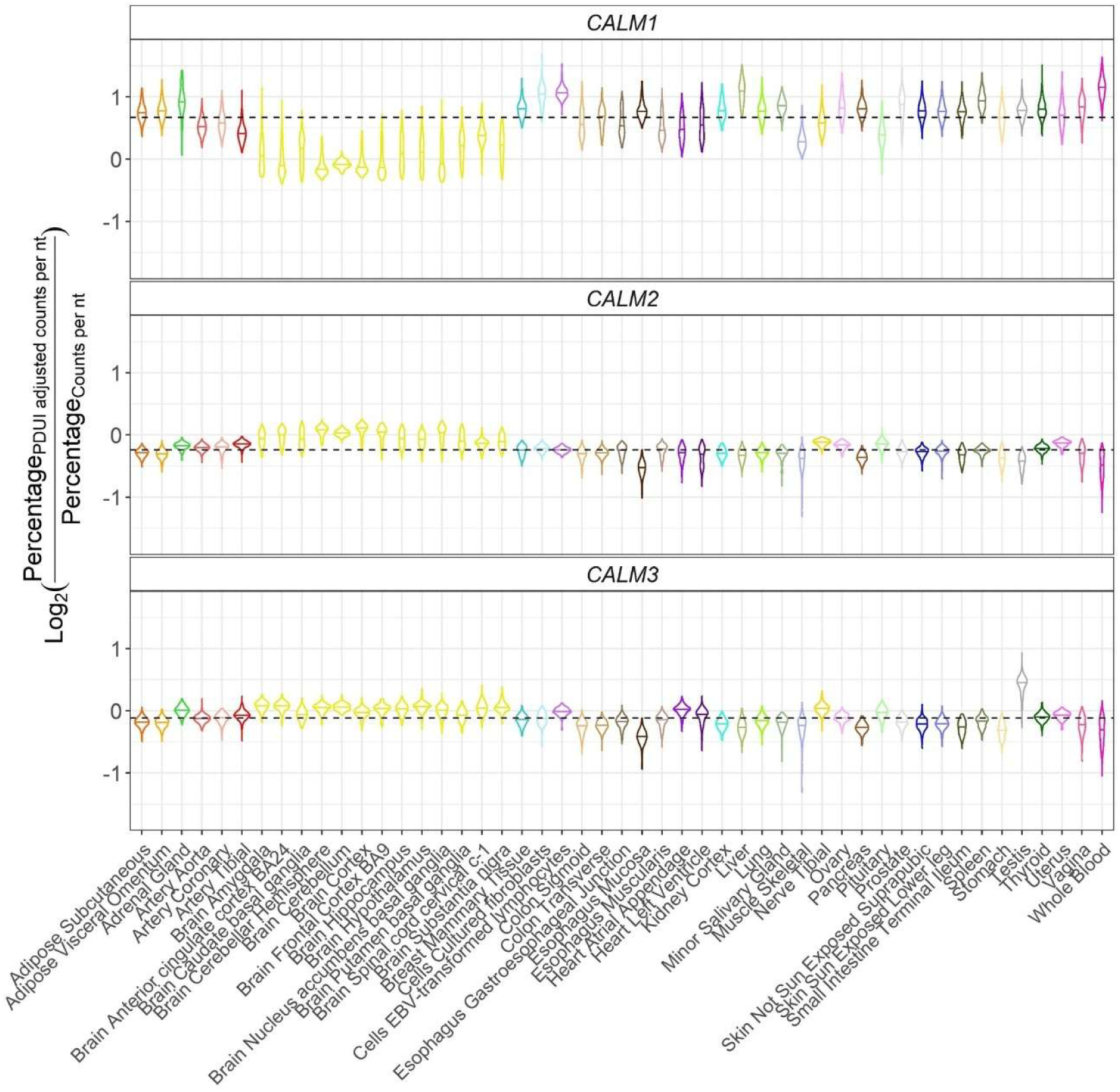
| Log2 transformed fold changes between the percentage of counts per gene measured as “PDUI adjusted counts per nt” and “counts per nt”. Violin plot of the log_2_ transformed fold changes between percentage of counts measured as “PDUI adjusted counts per nt” and “counts per nt per tissue among *CALM1, CALM2, and CALM3.* The horizontal line within each violin corresponds to the median fold change between the two measures for the particular tissue. The color codes indicate tissue of origin and are adopted from the Genotype-Expression Tissue (GTEx) project. The horizontal dashed lines in each of the plots shows the median of medians among the different tissues.

